# The risk of myocardial ischemia in patients with Kawasaki Disease: Insights from patient-specific simulations of coronary hemodynamics

**DOI:** 10.1101/2022.09.08.22279654

**Authors:** Karthik Menon, Jongmin Seo, Andrew M. Kahn, Jane C. Burns, Alison L. Marsden

## Abstract

**Background:** Pediatric patients with aneurysms due to Kawasaki disease require life-long and uninterrupted cardiology follow-up. Current AHA guidelines for risk stratification and long-term management are based primarily on maximal coronary artery luminal dimensions, normalized as Z-scores. Hemodynamic and functional significance of coronary arteries aneurysms on myocardial ischemic risk is not well studied.

**Methods:** We retrospectively studied a cohort of 15 patients who underwent coronary CT angiography imaging. We constructed patient-specific anatomic models from CT images and performed computational hemodynamic simulations incorporating pulsatile flow and deformable arterial walls. Simulation parameters were tuned to match patient-specific arterial pressure, ejection fraction, and cardiac output. From simulation results, we evaluated hemodynamic iscmemic risk metrics, including fractional flow reserve, wall shear stress, and residence time, in 153 coronary arteries.

**Results:** Fractional flow reserve (FFR) showed a weak correlation with aneurysm Z-scores. The slope of FFR significantly increased distal to the lumen narrowing at the end of aneurysms. Ischemic risk does not correlate well with aneurysm diameter measured by Z-score, but correlates much better with the ratio of maximum lumen diameter within aneurysms to minimum lumen diameter distal to aneurysms. Wall shear stress also correlates better with this diameter ratio, while residence can be stratified via Z-score.

**Conclusions:** Z-score alone is not a good indicator of abnormal FFR. Although FFR immediately distal to aneurysms is not critical, it starts to drop more rapidly distal to aneurysms and can therefore present an elevated risk of myocardial ischemia. Maximum-to-minimum lumen diameter is a good proxy for ischemic risk.

## I. Introduction

Kawasaki disease (KD) is an acute, self-limiting vasculitis of unknown etiology that occurs predominantly in children under 5 years of age [3, 23, 19]. KD is the most common cause of acquired heart disease in children in developed countries and the leading cause of acquired heart disease in children [19]. About 25% of KD patients develop coronary artery aneurysms or dilation without timely initiation of treatment (i.e. intravenous immunoglobulin), which may lead to thrombosis and sub-sequently myocardial infarction (MI), sudden death, or ischemic heart disease. Patients with giant aneurysms are at particularly high risk for coronary thrombosis, and require life-long cardiology follow-up. Current American Heart Association (AHA) recommendations for risk stratification and long-term management are based purely on maximum luminal diameter, in terms of Z-score or absolute dimension [19]. However, a better understanding of the functional significance of aneurysms in leading to adverse long-term outcomes requires detailed analyses of blood flow patterns as well as their role in the initiation of thrombosis. This is not available through standard imaging techniques and routine anatomical measurements.

The functional significance of coronary artery abnormalities in promoting ischemic risk can be assessed using the fractional flow reserve (FFR)[25, 26], which measures the flow reserve in arteries with disease compared to a hypothetical normal coronary artery under the same conditions. FFR is the gold standard for determining the ischemia-causing potential of atherosclerotic stenosis in adults, with FFR < 0.75 predominantly being used to indicate significant risk. However, FFR has not been as widely used to investigate ischemic risk in pediatric KD patients with coronary artery aneurysms. Ogawa *et al*. [24] performed a study to establish the effectiveness of FFR in assessing functional severity of coronary stenosis in pediatric patients with KD, and showed that a similar cutoff as that used in adults (FFR < 0.75) reliably indicates myocardial ischemic risk. Murakami and Tanaka [21] used coronary angiography to investigated the pressure drop caused by abnormal hemodynamics in aneurysms cased by KD and discussed integration of FFR as an additional method for risk stratification of KD patients. They showed that although abnormal hemodynamics in aneurysms did not lead to functionally significant pressure drop below the critical FFR threshold, there were statistically significant differences in FFR distal to giant aneurysms compared that distal to non-giant aneurysms. However, the hemodynamics and its role in pressure drop and functional significance of coronary artery aneurysms is still poorly understood.

Patient-specific cardiovascular modeling and flow simulation is an emerging technique which provides detailed quantitative estimates of local hemodynamics associated with the progression of cardiovascular diseases [15, 18, 28]. Computational simulations are being used to non-invasively estimate FFR, as in the FDA-approved FFTCT[36], and have proven to reduce unnecessary angiograms [20]. The validity of cardiovascular simulation tools has been examined and has shown good agreement with physical experimental counterparts [17] in cross-institutional studies [35] as well as invasive measurements [7]. Moreover, hemodynamic metrics obtained from cardiovascular simulations have been employed to study thrombosis risk due to coronary aneurysms in KD patients [11, 32, 33, 12]. These studies, using retrospective patient-specific simulations of coronary hemodynamics, showed that the hemodynamics quantities including low wall shear stress and residence time provide a better predictor of thrombosis risk than purely anatomical measures of aneurysm size.

In our work, we used patient-specific simulations of coronary hemodynamics to retrospectively compute FFR in 153 coronary arteries in 15 patients with KD. We performed simulations in anatomical models reconstructed from coronary computed tomography (CT) images, and evaluated FFR in virtual hyperemic conditions modeled by reduced coronary microvascular resistance. Specifically, we focused on the characterization of FFR in the region near the coronary artery aneurysms, where abnormal hemodynamics are expected due to enlargement and narrowing of vessel.

Our study is the first to non-invasively assess FFR in pediatric patients with KD, and investigate the relationship between aneurysm geometry and FFR as well as other hemodynamic metrics including time-averaged wall shear stress (TAWSS) and residence time (RT). We describe our methods for computational hemodynamics analysis and patient data acquisition in section II. The results on FFR and other hemodynamic metrics in the coronary artery aneurysms, and their correlation with aneurysm size and shape is reported in section III. Finally, we discuss the significance and clinical implications out of this research in section IV.

## II. Methods

### i. Patient Data

We retrospectively studied a cohort of 15 patients who underwent coronary CTA imaging for suspected KD-related coronary artery aneurysms (CAAs) at three centers – Lucile Packard Children’s Hospital at Stanford, CA, USA; University of California San Diego School of Medicine, La Jolla, CA, USA; and Nippon Medical School Hospital, Tokyo, Japan. This study was approved by the Institutional Review Board at Stanford University, University of California San Diego and Nippon Medical School. The inclusion criteria for this study were the availability of CTA images of sufficient resolution to identify CAAs and re-construct the 3D coronary anatomy, as well as clinical measurements of heart rate, blood pressure, cardiac output and ejection fraction measured at the time of the CTA. Cases with occlusions were not included in the analysis.

A summary of the patient cohort’s demographics, cardiac function and CAA sizes is provided in table 1. The CAA sizes were quantified using the Z-score metric of Dallaire & Dahdah [6], which is commonly used in clinical practice [19]. The cohort included a range of patient ages, from 3 to 59 years, as well as a range of patient sizes, measured using body surface area [13], from 0.58 to 2.1 m^2^. It included aneurysms with Z-scores in the range -1.61 to 28.12, and corresponding aneurysm diameters in the range 2.7 mm to 14.4 mm. Figure 1 shows an overview of the reconstructed coronary anatomies, highlighting a variety of aneurysm sizes as well as shapes.

**Table 1:** Summary of patient data. ID: Patient ID. Age range: Age range at CTA acquisition (exact age not reported to ensure anonymization). BSA: Body surface area. HR: Heart rate. BP: Blood pressure (systolic/diastolic). CO: Cardiac output. EF: Ejection fraction. LAD: Left anterior descending artery. LCX: Left circumflex artery. RCA: Right coronary artery. LM: Left main coronary artery. CAA: Coronary artery aneurysm.

| ID | Sex | Age range<br>[yrs] | BSA<br>[m<sup>2</sup>] | HR<br>[bpm] | BP<br>[mmHg] | CO<br>[L/min] | EF<br>[%] | CAA sizes<br>[Z-score] |
| --- | --- | --- | --- | --- | --- | --- | --- | --- |
| 1 | M | 6-10 | 1.16 | 59 | 105/52 | 3.4 | 60 | LAD: 8.0, LCX: 6.1, RCA: 8.6 |
| 2 | M | 26-30 | 2.09 | 70 | 98/61 | 4.2 | 60 | LAD: 6.7, LCX: 6.4, RCA: 10.06 |
| 3 | M | 21-25 | 1.70 | 42 | 123/81 | 3.8 | 60 | LAD: 3.4, LCX: 2.7, RCA: 3.4 |
| 4 | M | 11-15 | 1.21 | 56 | 82/43 | 2.8 | 62 | LAD: 5.2, RCA: 3.9 |
| 5 | M | 11-15 | 1.79 | 59 | 108/67 | 4.9 | 60 | LAD: 8.2, LCX: 6.5, RCA: 9.9 |
| 6 | M | 16-20 | 1.50 | 76 | 88/56 | 4.4 | 57 | LAD: 5.1, RCA: 4.67 |
| 7 | M | <= 5 | 0.76 | 105 | 90/55 | 4.3 | 60 | LAD: 8.8, LCX: 8.1 |
| 8 | M | 56-60 | 2.10 | 60 | 140/82 | 5.5 | 60 | LAD: 7.0, RCA: 9.3 |
| 9 | M | 11-15 | 1.10 | 71 | 76/44 | 4.2 | 71 | LAD: 13.8, LCX: 7.1, RCA: 14.4 |
| 10 | M | 16-20 | 1.92 | 74 | 127/78 | 5.2 | 65 | LAD: 3.1, LCX: 2.6, RCA: 2.7 |
| 11 | M | <= 5 | 0.58 | 93 | 94/56 | 2.4 | 62 | LAD: 9.6, LCX: 4.4, RCA: 5.4 |
| 12 | M | 16-20 | 1.50 | 113 | 109/62 | 6.8 | 67 | LM: 7.8, RCA: 6.4 |
| 13 | N/A | N/A | 0.95 | 78 | 106/48 | 3.8 | 51 | LAD: 2.32, LCX: 2.44, RCA: 2.0 |
| 14 | M | 36-40 | 1.70 | 78 | 109/58 | 4.9 | 69 | LM: 4.4, LCX: 7.9, RCA: 3.5 |
| 15 | M | 21-25 | 1.70 | 69 | 121/76 | 7.3 | 66 | LAD: 9.7, LCX: 3.3, RCA: 4.0 |

**Figure 1:**
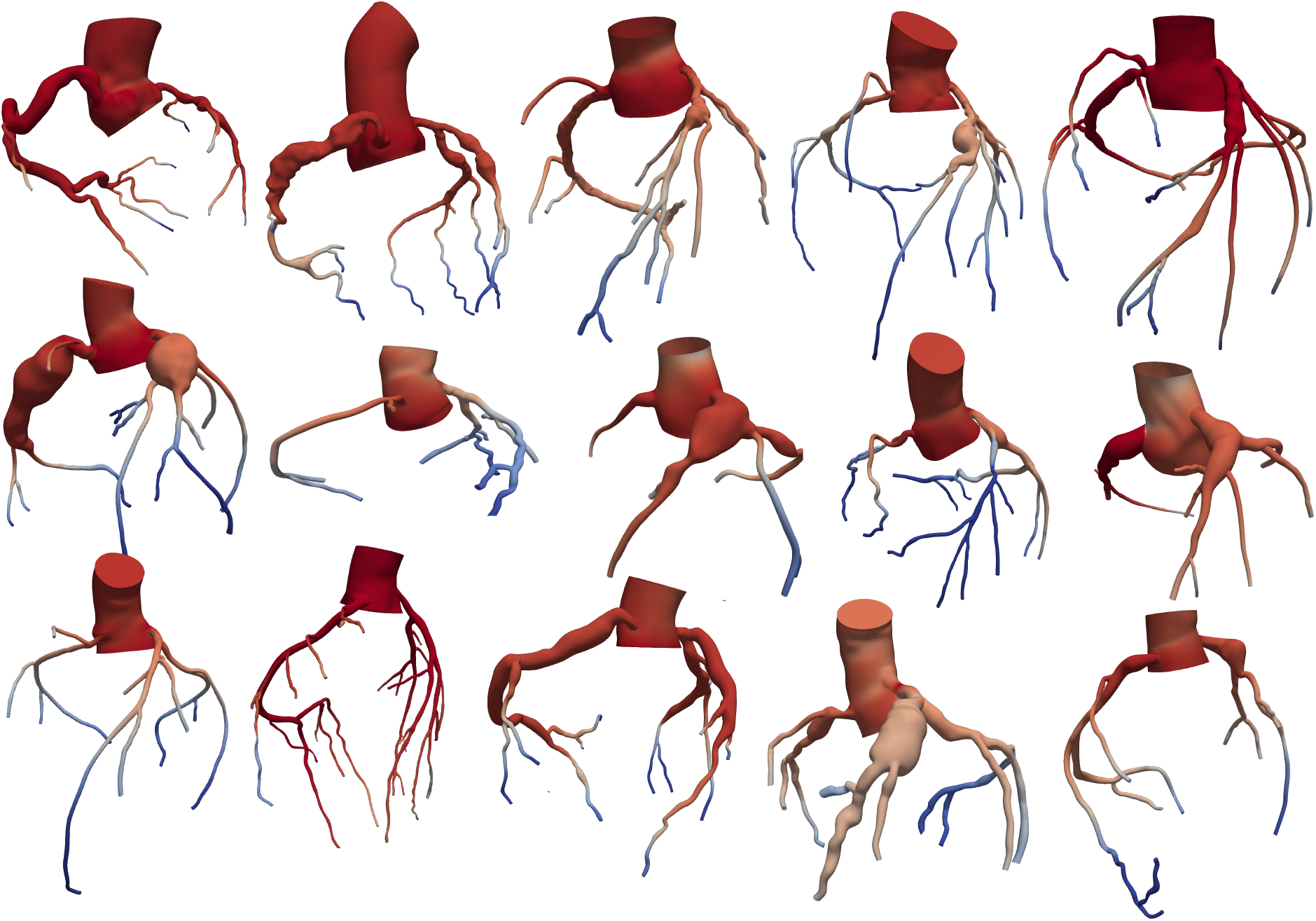
Overview of the 15 patient-specific coronary anatomy models with colors qualitatively indicating average pressure.

### ii. Patient-Specific Modeling and Simulations

The segmentation of coronary arteries from CT angiography images, construction of 3D anatomical models, and coronary flow simulations were performed using the open-source *SimVascular* software [38]. The modeling pipeline includes tools for tracing vessel centerlines, segmenting vessel lumen boundaries along the center-lines, and lofting the segmentations into a 3D anatomical model. Flow simulations were carried out using a stabilized finite element method [14], and finite element meshes were created using the open-source *TetGen* package [34], which is included with *SimVascular*. In our simulations, the vessel wall is compliant and deforms in response to the pressure and flow inside the lumen. This is enabled via the coupled momentum method [9]. The simulations yielded time-resolved blood velocity and pressure fields throughout the 3D coronary model.

For all cases simulated, blood was assumed to be a Newtonian fluid with viscosity 0.04 dynes/cm^2^ and density 1.06 g/cm^3^. Material properties for the artery walls were selected for each vessel based on its size and assigned values were based on our previous measurements as well as literature data. The elastic modulus for the aorta was set as 0.25 MPa [11] and that for coronary arteries was 1.15 MPa. [10, 5, 30]. Wall thicknesses were assigned from published radius-thickness ratios for the aorta [5] and morphometric data for the coronary arteries [27].

Boundary conditions for the blood flow simulations at inlets and outlets on the aorta and coronaries were enforced using a closed-loop lumped parameter network (LPN) model [16, 31, 37] shown in figure 2. This is a simplified and efficient representation of the coupling between the coronary/aortic outflow and the downstream vasculature and systemic circulation which brings blood back to the aortic inlet. It included specific features to model the resistance and capacitance of distal vasculature, the effect of the four heart chambers, and also captures the intramyocardial pressure experienced by coronary arteries that causes the out-of-phase flow behavior with respect to the cardiac cycle [15]. These parameters were tuned using an automated surrogate-based optimization framework [37] so that the resulting simulations reproduce the patient-specific clinically measured heart rate, aortic pressure, cardiac output and ejection fraction. The target coronary-systemic flow split was set at 4% based on in-vivo magnetic resonance velocity measurements [2] and we used a morphometric and physiological flow distribution amongst coronary arteries based on the modified Murray’s law [22, 40]. The automated parameter tuning resulted in good agreement between the simulations and the clinically measured targets. The average difference between simulation outputs and clinical measurements was 6.2% for systolic pressure, 7.4% for diastolic pressure, 4.2% for cardiac output, and 3.2% for ejection fraction.

**Figure 2:**
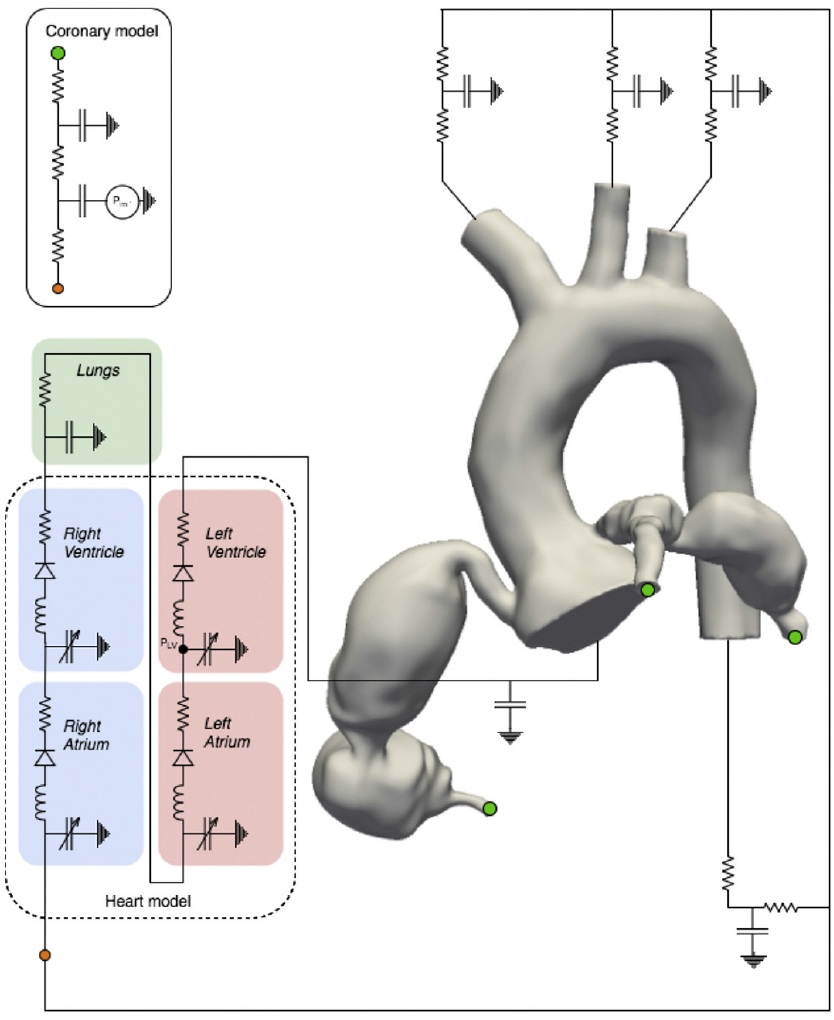
Simulation setup showing a three-dimensional patient-specific coronary artery model with a closed-loop circulation network.

Finally, once the boundary conditions were tuned to match patient-specific targets (measured at resting conditions), we performed simulations of the coronary flow in hyperemic conditions in order to replicate the effect of stress testing under adenosine. To simulate hyperemia, we reduced the resistance distal to coronary arteries (after they were tuned to match patient-specific targets at rest) to 0.24 times their values at the resting state. This is in line with the findings of Wilson et al. [39] who measured hyperemic coronary resistance with intravenous administration of adenosine, and also other widely used computational methods for simulating stress testing such as in FFR_CT_ [36].

### iii. Hemodynamic and Anatomical Metrics and Analysis

The analysis of data was performed on a vessel-by-vessel basis, and involved 153 coronary arteries from the 15 patient cohort. The main hemodynamic quantity of interest in this work is the fractional flow reserve (FFR) defined as the ratio of maximal blood flow to the myocardium through a diseased coronary artery to the theoretical maximal flow if the artery was normal [25]. In hyperemic conditions, FFR is measured at any point along a coronary artery as the mean pressure at that point divided by the mean aortic pressure. In adults with suspected coronary artery disease, FFR has been shown to accurately assess functionally significant ischemic risk and FFR values below 0.8 or 0.75 are generally considered critical [25, 26]. However, its use in pediatric patients is not as common, although past work suggests that similar FFR thresholds as those used in adult clinical practice are also useful for risk assessment in pediatrics [24].

We also analyzed the rate at which FFR drops along the length of each vessel, i.e. the slope of FFR. Mathematically, this was calculated using the gradient of FFR, *d*(FFR)/*dx*, where *x* is the length along the vessel. While FFR measures the fraction of aortic pressure available at any location along the artery, the slope of FFR measures how fast this pressure drops (as a fraction of the aortic pressure) as blood flows along the vessel. So if FFR itself is not critically low at a particular location in the artery, a steep slope of FFR indicates that FFR is dropping rapidly along the artery and can approach critical values at more distal locations. In this study, we particularly focused on the change in the slope (gradient) of FFR on either side of the lumen narrowing that occurs at the distal portion of each aneurysm. This highlights the effect of the distal lumen narrowing on increasing/decreasing the rate of pressure drop along the artery. We represent this change in slope by

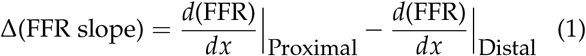

Other important hemodynamic quantities of interest included the time-average wall-shear stress (TAWSS) on vessel walls and the residence time (RT), both of which are related to ischemic risk via thrombus formation. The former is a measure of average blood flow-induced shear on the interior of vessel walls, which we measured using blood flow velocity vectors near the vessel walls obtained from patient-specific simulations. Abnormal wall-shear stress has been linked to thrombus formation in previous experiments that have studied the effect of pathological flow conditions on endothelial cell function [41, 4]. The latter quantifies the average time a fluid (blood) particle stays within a specific region of the model. Static or recirculating blood flow causes regions of high residence time, which when combined with low TAWSS has been associated with thrombosis [29, 1, 11]. Residence time was calculated by solving an advection equation using a finite element method [8]. This was computed within CAAs as a measure of thrombosis risk, and we report residence time in terms of the number of cardiac cycles a parcel of blood spends in the region of interest.

The above hemodynamic quantities of interest were analyzed with respect to two specific anatomical features – the Z-score calculated from vessel lumen diameters [6] and the ratio of the maximum-to-minimum lumen diameter for each aneurysm. The lumen diameters for both these metrics were calculated from the three-dimensional patient-specific anatomical models. We measured the maximum lumen diameter within the aneurysm and measured the minimum diameter immediately distal to the aneurysm, in order to isolate the hemodynamic effect of the lumen narrowing at the distal end of each aneurysm. For analyses across aneurysm sizes, we categorized the vessels in the cohort into Z-score categories of *Z <* 2 (no aneurysm), 2 *≤ Z <* 7.5 (small aneurysm), 7.5 *≤ Z <* 15 (medium aneurysm), *Z ≥* 15 (giant aneurysm). Statistical significance was assessed using non-parametric Wilcoxon rank-sum and signed-rank tests and degree of correlation is measured using the Pearson correlation coefficient (*r*_*P*_).

## III. Results

### i. FFR versus Z-score

Our results indicate that FFR measured distal to aneurysms is not strongly dependent on aneurysm diameter. This is evident from the vessel-by-vessel analysis of the current data set (figure 3) as well as individual patient-specific models (figure 4). Figure 3(a) shows the distribution of FFR for all vessels in each of the four Z-score categories mentioned in section iii. The median FFR distal to each aneurysm is non-critical for all categories, i.e. FFR *>* 0.8. Although giant aneurysms have lowest FFR values, the other categories are statistically indistinguishable. There is only a weak correlation with Z-score for the entire data set of vessels (*r*_*P*_ = *−*0.47).

**Figure 3:**
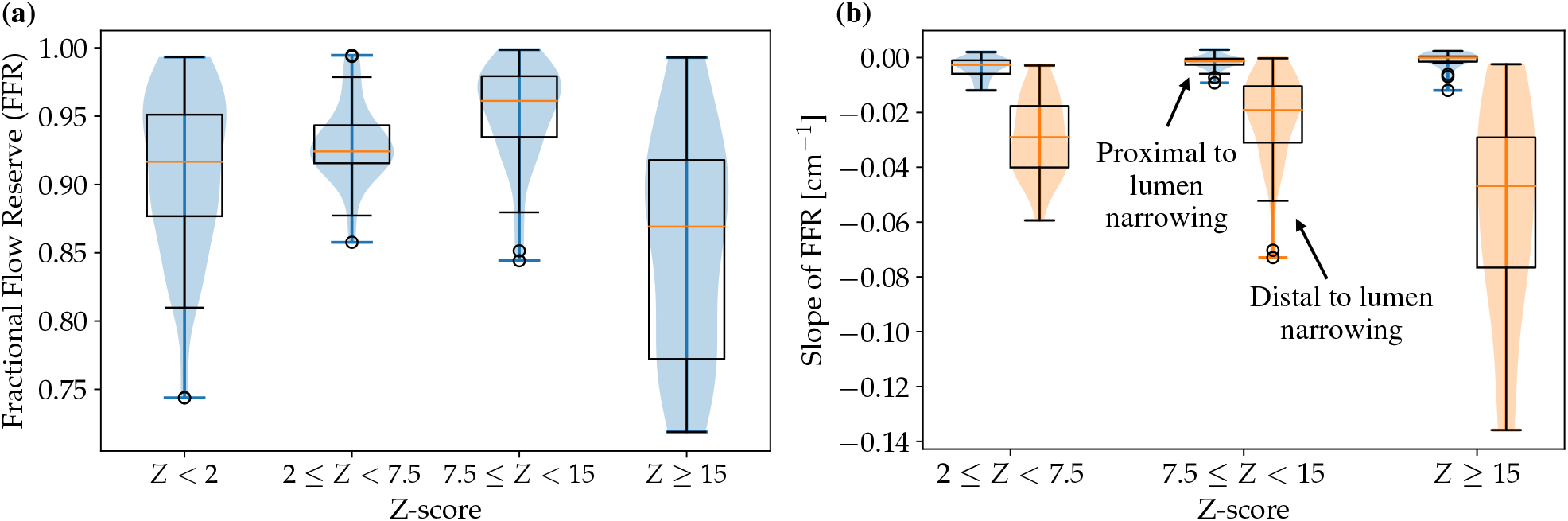
(a) Fractional flow reserve (FFR) distal to the aneurysm for all vessels in the cohort categorized into the four Z-score categories. (b) Difference in slope of FFR from proximal to distal of the lumen narrowing that occurs at the end of each aneurysm. Note: For each category, the box encloses the first to third quartile and whiskers represent 1.5 times the inter-quartile range. Circles represent outliers.

**Figure 4:**
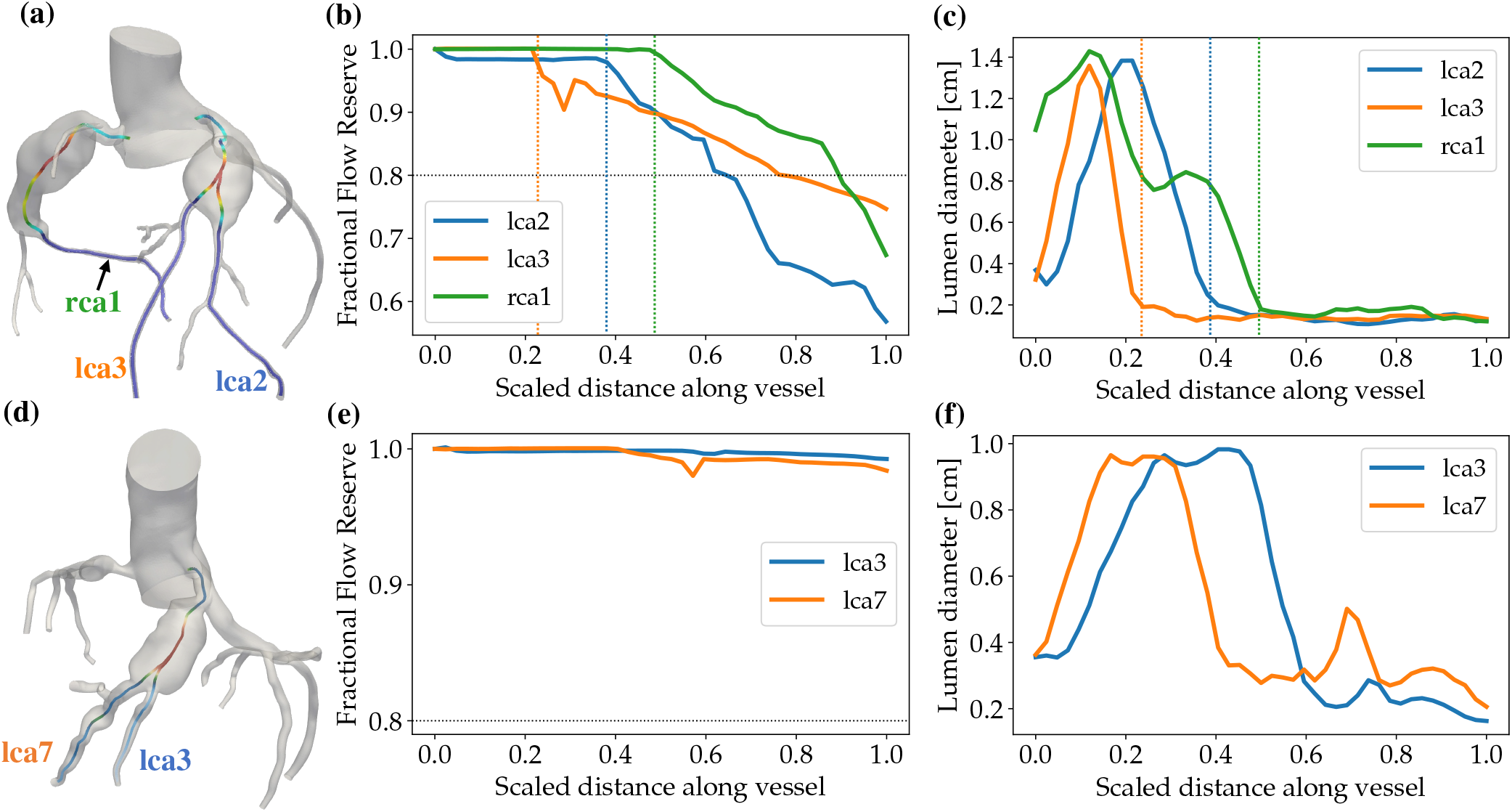
Comparison of FFR between two patients’ cases with similar Z-scores. (a),(d): Coronary artery anatomies with lumen centerlines of selected vessels highlighted. Centerline color qualitatively represents lumen diameter. (b),(e): FFR along the length of selected arteries. (c),(f): Lumen diameter along the length of selected arteries. In (b), (c), (e), (f), the X-axis is the lumen centerlines shown in (a) and (d), and the distance along the vessel is scaled with the total length of the selected vessel. The vertical lines in (b) and (c) highlight the locations of steepening FFR slope, which coincides with the aneurysm lumen narrowing.

We also find that the presence of CAAs changes the rate at which FFR drops along the length of each vessel. For each CAA Z-score category, figure 3(b) shows that the rate of FFR drop (which is negative because FFR reduces along the vessel) is faster, i.e. more negative, distal to the lumen narrowing that occurs at the distal end of each aneurysm. FFR drops much more rapidly distal to aneurysms for all aneurysm sizes (*p <* 10^*−*6^ for each Z-score category). Although the magnitude of this change is largest for giant aneurysms, there is no clear trend with respect to Z-score for small and moderate aneurysms.

The fact that Z-score alone does not predict ischemic risk can be further re-inforced by analyzing individual patient-specific models. For example, the two sample patient-specific models shown in figure 4 have aneurysms that correspond to similar Z-scores of 28 (LAD) and 24.4 (RCA) for the case shown in figure 4(a), and 23.2 (LAD) for the case in figure 4(d). However, FFR computed along the length of the vessel show different results. FFR for the case in figure 4(b) is below the value indicating critical ischemic risk, i.e. FFR *<* 0.80, whereas FFR for the case in figure 4(e) is above 0.95. Furthermore, these two cases show very different rates of FFR drop. Figure 4(b) shows a more rapid drop in FFR along the vessels compared to figure 4(e). It is also clear from figure 4(b) that the rapid drop in FFR begins at specific locations along the artery, which we have indicated using vertical dotted lines. By highlighting the corresponding locations in figure 4(c), we see that these locations at which the slope of FFR becomes steeper coincide with the locations where the lumen diameter reduces at the distal end of the aneurysm.

### ii. FFR versus maximum-to-minimum lumen diameter ratio

Although FFR value is not well predicted by the aneurysm diameter itself, the degree of lumen narrowing distal to the aneurysm is a better indicator of abnormal FFR for all vessels with measurable aneurysms (*Z ≥* 2). Figure 5(a) shows that the ratio of maximum-to-minimum lumen diameter close to the distal lumen narrowing for each aneurysm correlates inversely with FFR. Hence, more drastic lumen narrowing distal to aneurysms leads to lower FFR distal to aneurysms. Crucially, FFR correlates significantly better with lumen diameter ratio, with *r*_*P*_ = *−*0.64 (*p ≈* 10^*−*12^), compared with its correlation with Z-score (*r*_*P*_ = *−*0.47; *p≈*10^*−*6^). The current data indicates a trend between FFR and the lumen narrowing given by,

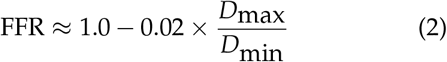

where *D*_max_/*D*_min_ is the maximum-to-minimum lumen diameter ratio.

**Figure 5:**
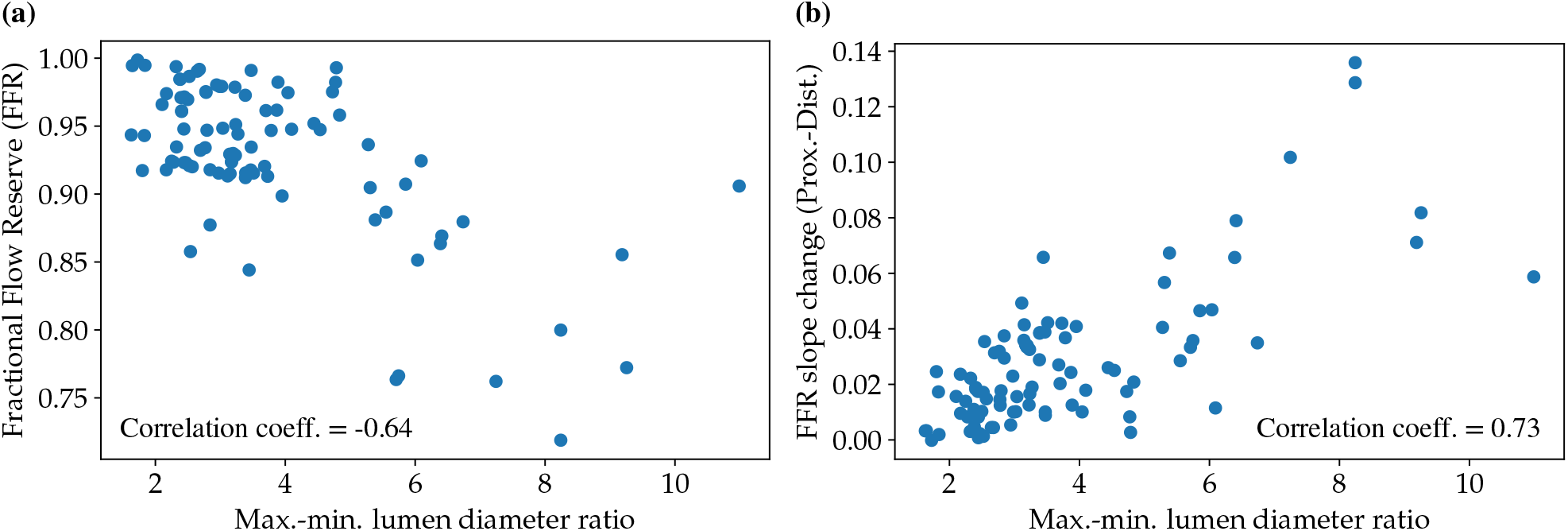
(a) FFR versus ratio of maximum to minimum lumen diameter for all CAA. (b) Change in FFR slope, Δ(FFR slope), from proximal to distal of the lumen narrowing that occurs at the end of each aneurysm, versus ratio of maximum to minimum lumen diameter for all CAA.

Moreover, figure 5(b) shows that the diameter ratio of the lumen narrowing in the distal portion of the aneurysms also correlates very well with the increase in the slope of FFR from the proximal to distal regions of this lumen narrowing. Therefore, FFR drops more rapidly distal to aneurysms which feature tighter distal lumen narrowing. The correlation coefficient of the change in FFR slope is *r*_*P*_ = 0.73 (*p ≈* 10^*−*16^) with respect to lumen diameter ratio, and only *r*_*P*_ = 0.54 (*p ≈* 10^*−*8^) with Z-score. The following linear fit can be obtained for the increase in FFR slope versus the lumen diameter ratio,

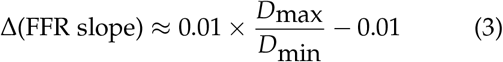

where Δ(FFR slope) is the increase in the slope of FFR from proximal to distal of the lumen narrowing measured in units of cm^−1^.

### iii. Lumen wall shear stress and blood residence time

Our patient-specific modeling framework allows the evaluation of TAWSS and RT. Previous work has demonstrated the occurrence of low TAWSS within aneurysms [11], which is supported by our findings in figure 6(a) for all categories of aneurysm size. As shown in figure 6(a), we find that TAWSS is lower inside aneurysms compared with the TAWSS measured distal to aneurysms. This difference in TAWSS outside and inside the aneurysm is most evident for giant aneurysms (*Z ≥* 15), but does not correlate well with Z-score across the full range of aneurysm sizes (*r*_*P*_ = 0.21; *p* = 0.04). It does, however, correlate better with both the ratio of maximum-to-minimum diameter, with correlation coefficient *r*_*P*_ = 0.51 (*p ≈* 10^*−*7^), as well as the change in FFR slope proximal and distal to the lumen narrowing (*r*_*P*_ = 0.59; *p ≈* 10^*−*10^), as seen in figure 6(b). On the other hand, we find that residence time of blood within the aneurysm shows a clear trend with aneurysm size, as seen in figures 6(c) and 6(d). For larger aneurysms, blood can persist for 2-4 cardiac cycles within an aneurysm and lead to enhanced clotting risk. The correlation with Z-score is moderate, with *r*_*P*_ = 0.52 (*p ≈* 10^*−*4^), and is slightly weaker with lumen diameter ratio (*r*_*P*_ = 0.47; *p* = 0.02).

**Figure 6:**
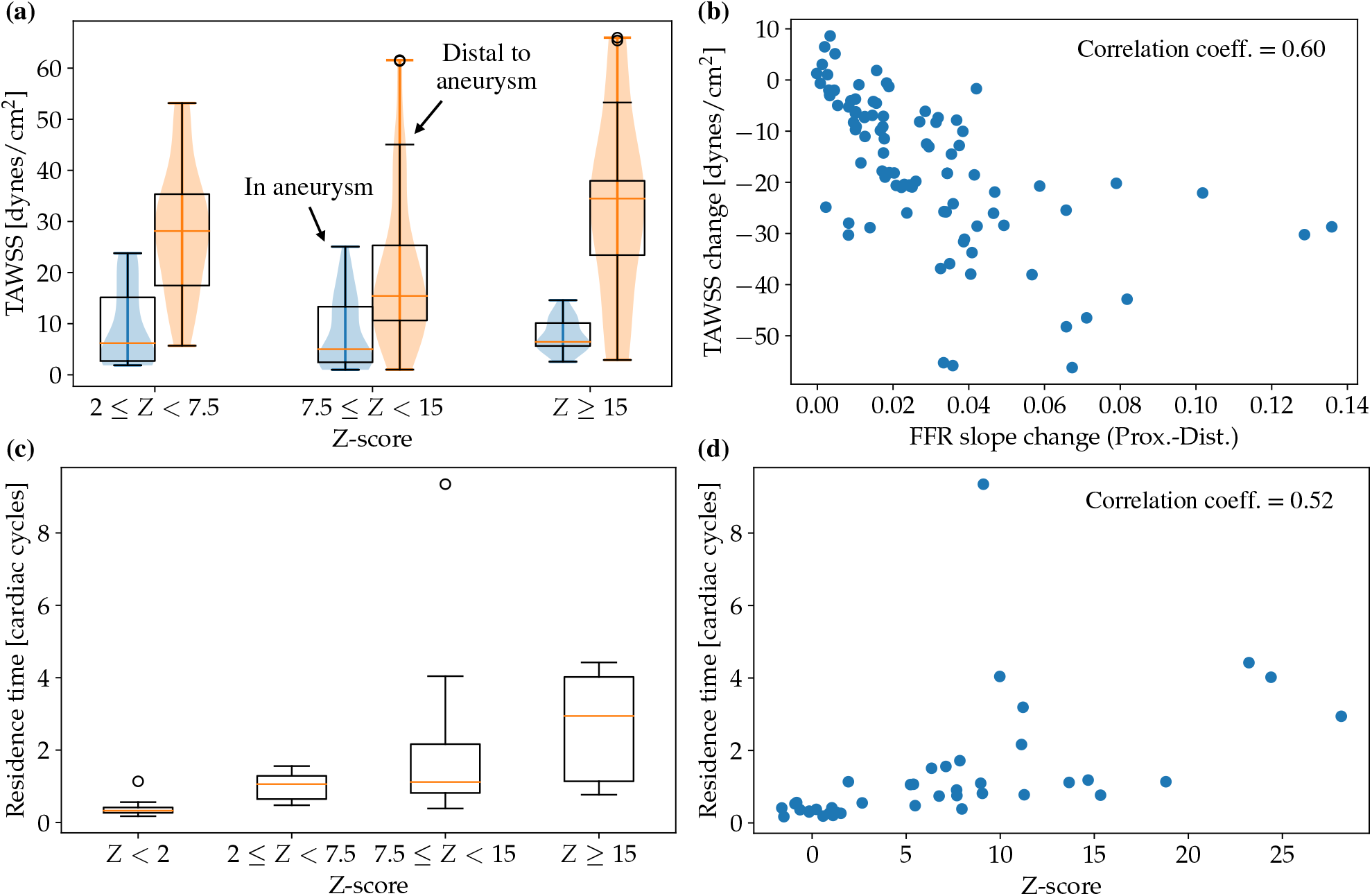
(a) Comparison of time-average wall shear stress (TAWSS) inside and outside the aneurysm for each Z-score category of CAA. (b) Difference between TAWSS inside and outside the aneurysm versus change in FFR slope. (c) Residence time in units of cardiac cycles for vessels in each Z-score category. (d) Residence time in terms of cardiac cycles versus Z-score. Note: For each category in (a) and (c), the box encloses the first to third quartile and whiskers represent 1.5 times the inter-quartile range. Circles in (a) and (c) represent outliers.

## IV. Discussion

To the authors’ knowledge, this paper is the largest patient and vessel cohort reported in a patient-specific computational study of coronary hemodynamics in Kawasaki disease. Although previous studies [33, 11] have highlighted the importance of hemodynamics in clinical risk stratification for KD treatment, the vessel-by-vessel analysis of 153 coronary arteries reported here reveals statistically significant insights that are missing from previous studies. Furthermore, while most previous work has focused on the risk of thrombosis in KD-induced aneurysms, this study is the first patient-specific computational investigation of ischemic risk, which is a relatively unexplored consequence of such coronary aneurysms.

### i. Functional Significance versus Aneurysm Geometry

An important finding from this study is that aneurysms can have a functionally significant effect on ischemic risk, but this is not evident from simply measuring FFR distal to aneurysms. We showed that although the median FFR measured 2 cm. distal to aneurysms is above critical thresholds for all categories of aneurysm size, FFR starts to drop rapidly downstream of the lumen narrowing distal to aneurysms. These findings are in-line with clinical invasive catheter-based measurements of FFR by Murakami and Tanaka [21] who found that FFR immediately distal to non-giant and giant aneurysms are non-critical, with giant aneurysms showing slightly lower FFR values. However, the rapid loss of pressure distal to aneurysms reported in this study can increases the risk of ischemia in distal portions of vessels and has not been investigated in previous work.

We showed that although the rate of FFR drop distal to aneurysms shows the greatest change distal to the largest aneurysms, it is statistically significant for all aneurysm sizes. In terms of ischemic risk, this finding suggests that otherwise mild stenotic lesions in distal portions of vessels could present more significant risks when they occur along with proximal aneurysms. This is because small pressure losses due to mild lesions can be exacerbated by the more rapid aneurysm-induced pressure loss – thus leading to critical ischemic risk.

Another important finding from this study is that aneurysms of similar diameter can feature significantly different flow conditions and present different clinical risks. This was also seen in previous studies focusing on thrombosis within aneurysms that result from KD [32, 33, 11]. We showed that FFR and the rapid drop of FFR distal to aneurysms are not a strong function of aneurysm diameter measured by Z-score, with correlation coefficient *r*_*P*_ = *−*0.47 and *r*_*P*_ = *−*0.54 respectively with Z-score. However, both these measures of functional ischemic risk correlate much more strongly with the degree of narrowing distal to aneurysms. The correlation coefficient of FFR with lumen diameter ratio is *r*_*P*_ = 0.64, and the change in FFR slope distal to aneurysms correlates with lumen diameter ratio with a correlation coefficient *r*_*P*_ = 0.73. This strong influence of lumen narrowing suggests a stenosis-like hemodynamic mechanism for pressure loss distal to aneurysms, which was also alluded to by Murakami and Tanaka[21].

While the above findings evaluate ischemic risk due to disturbed flow within aneurysms, we also evaluated residence time and wall-shear stress metrics that relate to the longer-term development of occlusive thrombosis within aneurysms. Previous work has correlated high residence time with thrombus formation [33, 11], and we showed that large Z-scores correlate moderately well with high residence time (*r*_*P*_ = 0.52) in the current dataset. The role of wall-shear stress in thrombus formation has been documented in previous experimental studies [41, 4] that have linked hemodynamic stimuli on endothelial cell function with blood coagulation. Here, we reinforced previous findings using a different patient cohort that wall-shear stress is lower within aneurysms [11]. Furthermore, we also showed that the lower wall-shear stress within aneurysms correlates much better with the degree of lumen narrowing (*r*_*P*_ = 0.51) than the Z-score of the aneurysm (*r*_*P*_ = 0.21). This highlights a possible interesting connection between the hemodynamics that cause FFR drop with that of thrombosis development within aneurysms.

### ii. Clinical Implications

One clinically significant outcome from this work is evidence that aneurysms can drive ischemic risk due to the direct influence of abnormal blood flow, not only due to the longer-term development of occlusion. Current notions of CAA ischemic risk are usually related to the thrombosis, luminal myofibroblastic proliferation, and sometimes atherosclerosis. These factors can lead to the development of coronary artery occlusion and myocardial ischemia in the long-term [19]. However, in this study we showed that the presence of aneurysms in coronary arteries can have negative near-term consequences measured by FFR, even in the absence of longer-term occlusion development.

Of great clinical importance is the relevance of the current findings to American Heart Association (AHA) treatment guidelines [19]. The primary clinical intervention for CAAs that result from KD is anti-coagulation therapy targeted at preventing thrombosis. Firstly, our findings indicate that treatment guidelines would benefit from considering near-term ischemic risk from abnormal blood flow in addition to longer-term thrombosis risk. Secondly, the treatment guidelines are based purely on aneurysm size and Z-score, but our study shows that Z-score alone is not a good predictor of ischemic or thrombosis risk. We show here that the lumen diameter ratio, or the degree of narrowing distal to an aneurysm, correlates better with measures of FFR for ischemic risk than Z-score. The importance of the lumen narrowing indicates a potential stenosis-like hemodynamic mechanism driving ischemic risk. This is in-line with previous studies that have highlighted the superiority of hemodynamics-driven risk stratification over anatomical risk stratification for KD-induced aneurysms [32, 33, 11] as well as for coronary artery disease [26, 36]. However, it is important to remark that since the lumen diameter ratio is a purely anatomy-based proxy for a hemodynamic effect, it can be easily incorporated into future treatment guidelines (when validated with more clinical data) even if they are based purely on non-invasive anatomical imaging.

Nevertheless, this work highlights potentially important hemodynamic metrics, such as the rate of FFR drop, which are not clinically accessible using non-invasive methods and can usually only be measured using simulations. The analysis of such hemodynamic drivers of disease risk, and the fact that clinical risks and flow patterns are not determined purely by aneurysm size, underscore the relevance of personalized computational tools in clinical assessment of functional risk.

### iii. Limitations and Future Work

Although this study includes a larger patient cohort than previous patient-specific computational studies, patients numbers would need to be higher to realistically inform widely-accepted clinical treatment standards and guidance. The number of patients was limited by the availability of CTA and ultrasound data with high enough quality to build and tune patient-specific models. Furthermore, availability of data on clinical outcomes and measurements would enhance the impact of these findings on clinical translation. We also did not consider the effect of multiple aneurysms and/or lesions on ischemic risk. This will be a subject of future study.

The use of FFR to measure ischemic risk is less common in pediatric populations than in adults, and there are no widely-used FFR thresholds to signify functional severity in children. The thresholds used here were based on the work of Ogawa et al. [24], who showed that FFR is a good measure of functional ischemic severity in children, with similar risk thresholds as those used in adults. However in leiu of more data, the use of more pediatric-specific metrics for ischemic risk and severity cut-offs would give this work better context.

This work opens several avenues for future research into the ischemic risk presented by coronary artery aneurysms in patients with KD. For one, the abnormal flow that dictates pressure loss could have more complex dependencies that have not been explored in this work. Although correlations with more established shape-related metrics were explored in this study, we found that they did not correlate better with FFR than the lumen diameter ratio reported. The use of virtual modeling tools would allow more rigorous investigations into the effect of aneurysm shape on hemodynamics and clinical outcomes.

## V. Conclusions

In this work, we used patient-specific computational simulations to retrospectively evaluate the risk of myocardial ischemia in patients with Kawasaki disease. We analyzed 15 patient-specific models consisting of 153 coronary arteries on a vessel-by-vessel basis and fractional flow reserve was used to estimate ischemic risk. We showed that although FFR immediately distal to aneurysms is not critical, it starts to drop more rapidly distal to aneurysms and can therefore present an elevated risk of myocardial ischemia. We showed that ischemic risk does not correlate well with aneurysm diameter measured by Z-score, but correlates much bettter with the ratio of maximum-to-minimum lumen diameter distal to aneurysms. This main takeaway from this study is that coronary artery aneurysms can potentially present a risk of myocardial ischemia even in the absence of long-term thrombosis and occlusion development.

## Data Availability

All data produced in the present study are available upon reasonable request to the authors.

## VI. Acknowledgements

We are grateful to the Hooper Family Trust for funding this study.

